# Bayesian network modeling of risk and prodromal markers of Parkinson’s disease

**DOI:** 10.1101/2022.05.18.22275239

**Authors:** Meemansa Sood, Ulrike Suenkel, Anna-Katharina von Thaler, Helena U. Zacharias, Kathrin Brockmann, Gerhard W. Eschweiler, Walter Maetzler, Daniela Berg, Holger Fröhlich, Sebastian Heinzel

## Abstract

Parkinson’s disease (PD) is characterized by a long prodromal phase with a multitude of markers indicating an increased PD risk prior to clinical diagnosis based on motor symptoms. Current PD prediction models do not consider interdependencies of single predictors, lack differentiation by subtypes of prodromal PD, and may be limited and potentially biased by confounding factors, unspecific assessment methods and restricted access to comprehensive marker data of prospective cohorts.

We used prospective data of 20 established risk and prodromal markers of PD in 1178 healthy, PD-free individuals and 24 incident PD cases collected longitudinally in the Tübingen evaluation of Risk factors for Early detection of NeuroDegeneration (TREND) study at 4 visits over up to 10 years. We employed artificial intelligence (AI) to learn and quantify PD marker interdependencies via a Bayesian network (BN) with uncertainty estimation using bootstrapping. The BN was employed to generate a synthetic cohort and individual marker profiles.

Robust interdependencies were observed for BN edges from age to subthreshold parkinsonism and urinary dysfunction, sex to substantia nigra hyperechogenicity, depression, non-smoking and to constipation; depression to symptomatic hypotension and excessive daytime somnolence; solvent exposure to cognitive deficits and to physical inactivity; and non-smoking to physical inactivity. Conversion to PD was interdependent with prior subthreshold parkinsonism, sex and substantia nigra hyperechogenicity. Several additional interdependencies with higher statistical uncertainty were identified. Synthetic subjects generated via the BN based representation of the TREND study were realistic as assessed through multiple comparison approaches of real and synthetic data.

Altogether our work demonstrates the potential of modern AI approaches (specifically BNs) in two ways: First, to model and understand interdependencies between PD risk and prodromal markers, which are so far not accounted for in PD prediction models. Second, the generative nature of BNs opens the door for facilitating data sharing in a legally compliant and privacy preserving manner.

## 1.0 Introduction

Parkinson’s disease (PD) is characterized by progressive neurodegeneration that has usually advanced for many years before PD is clinically diagnosed on the basis of its cardinal motor symptoms (1). In addition to old age, a multitude of risk markers, such as genetic factors, lifestyle, environmental factors, (comorbid) diseases (e.g., diabetes) as well as biomarkers (e.g., low plasma urate levels, hyperechogenicity of the substantia nigra) have been shown to indicate an increased risk of PD in prospective studies (2,3). Moreover, prodromal markers may already indicate early neurodegenerative processes that can ultimately lead to the clinical diagnosis of PD. Depression, autonomous dysfunction, REM-sleep behavior disorder (RBD), subtle motor signs and pathological dopaminergic imaging are among the most established prodromal markers (3–5). The International Parkinson and Movement Disorder Society (MDS) research criteria for prodromal PD (3,6) have been designed to review and continually update the predictive values of risk and prodromal markers of PD as indicated by the positive and negative likelihood ratio (LR+, LR-) calculated from a 2×2 table of prospective data: marker present/absent and incident PD diagnosis/healthy. Moreover, these criteria proposed a naïve Bayesian classifier approach for the calculation of the probability that an individual is in the prodromal phase of PD. With age providing an *a-priori* probability of prodromal PD as derived from epidemiological evidence (7), the individual profile of risk and prodromal markers, i.e. constellations of LR+ and LR-values, allows the calculation of an *a-posteriori* probability of prodromal PD (6). While the criteria have repeatedly been shown to be highly specific, sensitivity may depend upon marker selection, depth of assessment and time to PD diagnosis and possibly specific subtypes of prodromal PD (8, 9). While having the advantages of being both evidence-based as well as practical, several limitations and assumptions are inherent to this approach, that should be addressed to improve the accuracy of PD prediction. Most critically, statistical independence of risk markers and prodromal markers as well as age is assumed when using a naïve Bayesian classifier, which is most likely not fulfilled in reality. For example, many prodromal markers (including subtle motor deficits) increase in prevalence with advancing age irrespective of a future PD diagnosis, which may decrease their specificity for the prediction of PD in an age-dependent manner (10). Also, marker prevalence and their predictive value for PD may be sex-specific, e.g., as previously suggested for depression (10). Thus, the predictive value for PD as currently assigned to the presence, absence or borderline status of a particular marker, which may partially depend on age as well as constellations of the presence and absence of other markers in the profile of an individual. Marker co-occurrences may (partially) depend on e.g., methodological aspects of data collection and marker assessment, shared bio-pathological pathways and clinical comorbidity features. Such interdependencies can influence the actual predictive value of specific marker constellations. For instance, markers of different autonomous dysfunctions or depression and cognitive deficits and their predictive values might be interdependent for practical, statistical, biological and/or clinical reasons.

Evidence-based PD prediction approaches, such as the MDS research criteria for prodromal PD (3,6), may be limited in accuracy and validity due to restricted access to data and evidence obtained in prospective studies. Interdependencies, confounders and interactions of markers, demographic and cohort-specific variables, and details of the temporal changes of the predictive values of markers towards the clinical PD diagnosis, are often not considered or reported in detail in original publications (11). If reported, statistical methods often differ between publications complicating their integration into a common statistical framework of PD prediction. In this regard the MDS criteria offer a practical approach for considering published evidence of predictive markers of PD as often reported or available upon request. However, these LR values may be partly confounded (e.g., by age or other markers) and/or biased (e.g., by attrition) while temporal information is often missing and thus cannot be considered in PD prediction models (11). However, access to comprehensive data and evidence of predictive PD markers is often restricted due to legal and ethical considerations that prohibit the sharing of patient-level data.

The heterogeneity of PD in its clinical as well as in its prodromal phase may be partially explained by different subtypes of the disease (8, 9), e.g., subtypes differentiated by the site of initiation and progression of pathology (brain-first vs. body-first) (12,13). Such subtypes would likely also differ in risk and prodromal marker profiles (possibly cognitive deficits/depression in brain-first; autonomous dysfunction in body-first PD), and potentially in the temporal dynamics in the prodrome of PD. However, as comprehensive data of (major) prospective population-based cohorts is often not jointly accessible, early (prodromal) subtyping, predictive values of markers (and their interdependencies) by subtype is still largely restricted to highly selected and specific clinical populations such as RBD patients. Consequently, an evidence-based understanding of prodromal PD to improve PD prediction and aid the (subtype-specific) recruitment of future early intervention trials in prodromal PD is challenged by the unavoidable statistical biases of each clinical study due to predefined patient selection criteria.

Artificial Intelligence (AI) approaches, such as Bayesian networks (BNs) (14), may offer possible solutions to these challenges, as 1) interdependencies of markers can be modelled, 2) BNs can be used to realistically simulate prospective cohorts, which could – at least partially – help to overcome restrictions posed by data privacy, and 3) access to such synthetic, comprehensive (population-based) cohort data. Thereby, both the consideration of more generalizable evidence underlying PD prediction as well a more differentiated investigation and understanding of prodromal PD subtypes may be supported and possibly help to inform the design and recruitment for early intervention trials in prodromal PD.

The present study has two different aims: 1) to model a BN with the interdependencies between longitudinal data of risk and prodromal markers of PD and incident PD status of a large prospective cohort (TREND study), and 2) to demonstrate the feasibility of generating a sufficiently realistic synthetic cohort, which shares statistical patterns of the original data and could allow researchers to gain a better understanding of properties of the real data before formally applying for access to it.

## 2.0 Materials and Methods

### 2.1 Overview of the TREND study data

The TREND study is a prospective cohort study which has been conceptualized for the investigation of markers that may help to predict PD and/or Alzheimer’s disease (AD). The cohort is partly population-based and partly enriched with individuals with an increased PD/AD risk by selectively recruiting participants based on the presence of olfactory loss, depression, and/or possible RBD. Comprehensive assessments of risk and prodromal markers of neurodegeneration, and e.g., neurological, neuropsychiatric and quantitative motor testing as well as biosampling in 1,201 individuals (aged 50+ years at baseline), have been performed every two years (baseline in 2009/2010, follow-up 1 to 4; follow-up 5 is currently ongoing). For more information, visit https://www.trend-studie.de/english. The study was approved by the local ethics committee (Medical Faculty, University of Tübingen; 444/2019BO2). All participants provided written informed consent. Study data were collected and managed using REDCap electronic data capture tools hosted at University of Tübingen (9).

Cohort participants in part had a delayed inclusion in the study (at follow-up 1) and some participants missed single waves or dropped out of the study (retention rate at follow-up 4: 72.4%). Therefore, the number of individual visits instead of the wave number of the TREND study is considered in the present work. For some participants the duration between two visits may occasionally be longer than two years. After excluding individuals with PD or parkinsonism at visit 1, we included data of 1178 (98.08%) participants collected at four consecutive visits (Tables 1 and 2) as the number of individuals with five visits was substantially lower (n=545, 45.4%). Changes in sample size between visits as well as missingness of marker assessments per visit are shown in Tables 1 and 2. For the BN approach missingness both due to study drop-out (until visit 4) as well as due to missingness of marker assessment at single visits was considered for the imputation of data (see below and Supplementary material).

**Table 1.** Summary statistics of age and risk markers of PD-free individuals and incident PD cases at different visits, absolute and relative (%) frequencies of marker presence or median (inter-quartile range in brackets) are given unless specified otherwise. Sample sizes per visit are indicated. Missingness of marker data within a given visit is indicated, i.e. not considering longitudinal study dropout. Percentage values indicate the relative frequency of marker presence/absence/borderline/missingness within PD-free and incident PD groups, respectively, and within each visit. GBA, glucocerebrosidase; PD, Parkinson’s disease; SN, substantia nigra.

| Features | Category | Visit 1 | Visit 2 |  | Visit 3 |  | Visit 4 |  |
| --- | --- | --- | --- | --- | --- | --- | --- | --- |
|  |  | PD-free<br>(n=1178) | PD-free<br>(n=1070) | Incident<br>PD (n=6) | PD-free<br>(n=981) | Incident<br>PD (n=5) | PD-free<br>(n=880) | Incident<br>PD (n=5) |
| *Age at visit 1* |  | 63 (58, 68) | 65 (60, 70) | 74 (70, 74) | 67 (62, 72) | 75 (73, 76) | 69 (64, 74) | 70 (68, 71) |
| *Sex* | Male | 599 (51%) | 546 (51%) | 5 (83%) | 503 (51%) | 5 (100%) | 464 (53%) | 3 (60%) |
|  | Female | 579 (49%) | 524 (49%) | 1 (17%) | 478 (49%) | 0 (0%) | 416 (47%) | 2 (40%) |
| *PD family history* | No | 1010 (86%) | 917 (86%) | 3 (50%) | 840 (86%) | 4 (80%) | 751 (85%) | 4 (80%) |
|  | Yes | 168 (14%) | 153 (14%) | 3 (50%) | 141 (14%) | 1 (20%) | 129 (15%) | 1 (20%) |
| *Polygenic risk score* | Marker absent | 247 (21%) | 218 (20%) | 2 (33%) | 204 (21%) | 1 (20%) | 181 (21%) | 3 (60%) |
|  | Borderline | 500 (42%) | 459 (43%) | 2 (33%) | 420 (43%) | 2 (40%) | 380 (43%) | 1 (20%) |
|  | Marker present | 252 (21%) | 237 (22%) | 1 (17%) | 218 (22%) | 1 (20%) | 197 (22%) | 0 (0%) |
|  | Missing | 179 (15%) | 156 (15%) | 1 (17%) | 139 (14%) | 1 (20%) | 22 (14%) | 1 (20%) |
| *GBA mutation* | No | 1126 (96%) | 1021 (95%) | 6 (100%) | 938 (96%) | 2 (40%) | 845 (96%) | 4 (80 %) |
|  | Yes | 52 (4%) | 49 (5%) | 0 (0%) | 43 (4%) | 3 (60%) | 35 (4%) | 1 (20%) |
| *SN hyperechogenicity* | SN- | 839 (71%) | 771 (72%) | 1(17%) | 714 (73%) | 3 (60%) | 641 (73%) | 3 (60%) |
|  | SN+ | 210 (18%) | 191 (18%) | 5 (83%) | 178 (18%) | 2 (40%) | 168 (19%) | 2 (40%) |
|  | Missing | 129 (11%) | 108 (10%) | 0 (0%) | 89 (9%) | 0 (0%) | 71 (8%) | 0 (0%) |
| *Occupational pesticide exposure* | No | 878 (75%) | 855 (80%) | 1(17%) | 839 (86%) | 1 (20%) | 813 (92%) | 3 (60%) |
|  | Yes | 19 (2%) | 18 (2%) | 0 (0%) | 17 (2%) | 0 (0%) | 17 (2%) | 0 (0%) |
|  | Missing | 281 (24%) | 197 (18%) | 5 (83%) | 125 (13%) | 4 (80%) | 50 (6%) | 2 (40%) |
| *Occupational solvent exposure* | Yes | 774 (66%) | 753 (70%) | 1 (17%) | 739 (75%) | 1 (20%) | 714 (81%) | 2 (40%) |
|  | No | 129 (11%) | 125 (12%) | 0 (0%) | 122 (12%) | 0 (0%) | 117 (13%) | 1 (20%) |
|  | Missing | 275 (23%) | 192 (18%) | 5 (83%) | 120 (12%) | 4 (80%) | 49 (6%) | 2 (40%) |
| *Diabetes type II* | No | 1131 (96%) | 1015 (95%) | 5 (83%) | 922 (94%) | 5 (100%) | 826 (94%) | 4 (80%) |
|  | Yes | 47 (4%) | 55 (5%) | 1 (17%) | 59 (6%) | 0 (0%) | 54 (6%) | 1 (20%) |
|  | Missing | 0 (0%) | 0 (0%) | 0 (0%) | 0 (0%) | 0 (0%) | 0 (0%) | 0 (0%) |
| *Physical inactivity* | No | 542 (46%) | 343 (32%) | 1 (17%) | 754 (77%) | 4 (80%) | 701 (80%) | 3 (60%) |
|  | Yes | 142 (12%) | 98(9%) | 0 (0%) | 223 (23%) | 1 (20%) | 176 (20%) | 2 (40%) |
|  | Missing | 494 (42%) | 629 (59%) | 5 (83%) | 4 (0%) | 0 (0%) | 3 (0%) | 0 (0%) |
| *Non-smoking* | Marker absent | 535 (45%) | 478 (45%) | 2 (33%) | 438 (45%) | 3(60%) | 396 (45%) | 0 (0%) |
|  | Borderline | 533 (45%) | 509 (48%) | 4 (67%) | 470 (48%) | 2(40%) | 422 (48%) | 4 (80%) |
|  | Marker present | 109 (9%) | 83 (8%) | 0 (0%) | 72 (7%) | 0 (0%) | 61 (7%) | 1 (20%) |
|  | Missing | 1 (0.1%) | 0 (0%) | 0 (0%) | 1 (0.1%) | 0 (0%) | 1 (0.1%) | 0 (0%) |

**Table 2.**
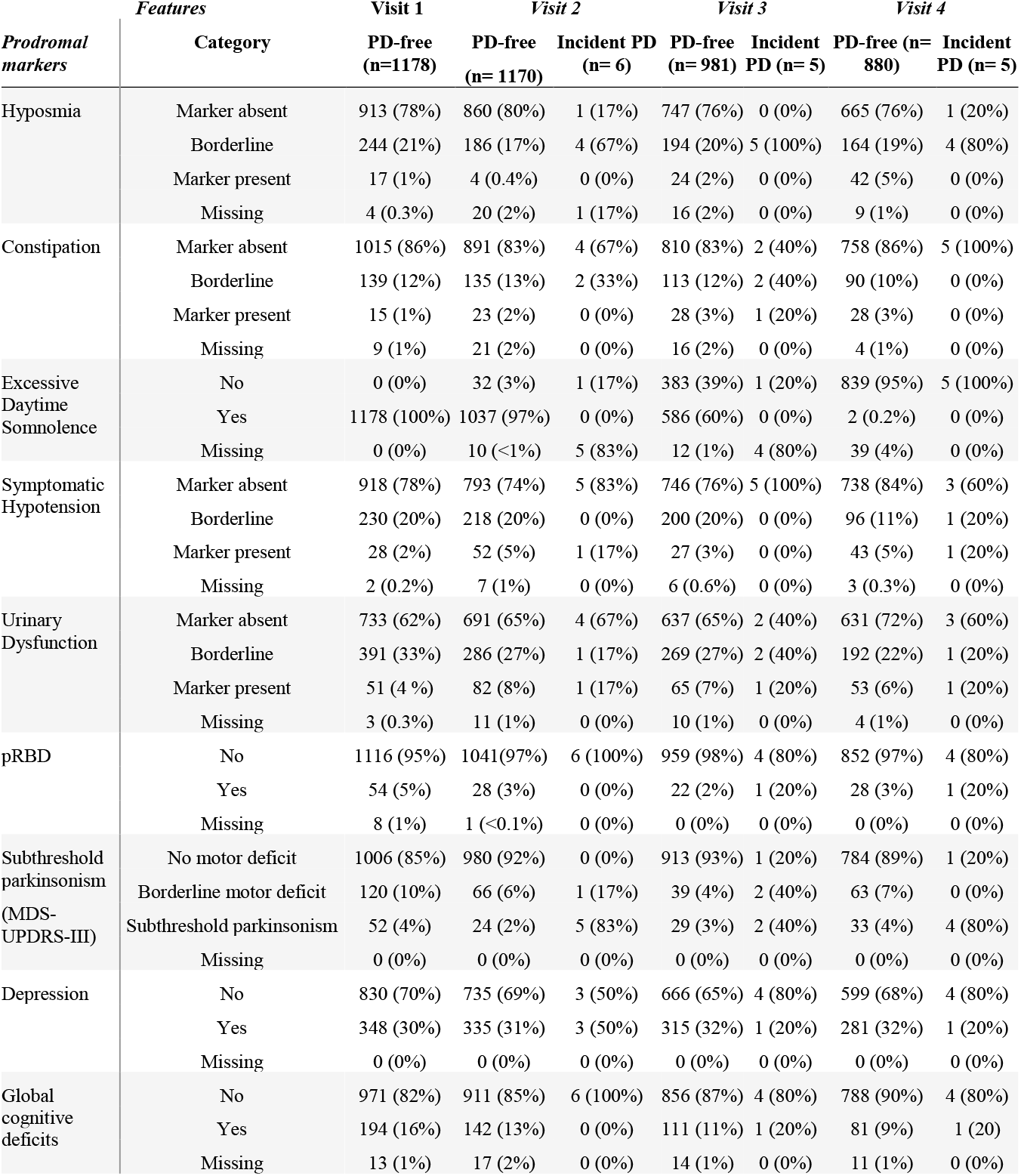
Summary statistics of prodromal markers of PD-free individuals and incident PD cases at different time points, absolute and relative (%) frequencies of marker presence or median (inter-quartile range in brackets) are given unless specified otherwise. Sample sizes per visit are indicated. Missingness of marker data within a given visit is indicated, i.e. not considering longitudinal study dropout. Percentage values indicate the relative frequency of marker presence/absence/borderline/missingness within PD-free and incident PD groups, respectively, and within each visit. MDS-UPDRS-III, MDS-sponsored Unified Parkinson’s Disease Rating Scale, motor part 3; PD, Parkinson’s disease, pRBD, possible REM sleep behavior disorder.

While all of these participants were PD-free at the first visit, in total n=24 incident PD cases were clinically diagnosed at follow-up based on UKBB and MDS diagnostic criteria (15). The visit at which the conversion to PD occurred was considered, and descriptive statistics of PD-free individuals and incident PD cases regarding demographic factors and risk and prodromal markers of PD are shown in Tables 1 and 2.

### 2.2 Bayesian Networks based approach

We propose a BN based approach (11) to model the interdependencies between different risk and prodromal markers of PD and their longitudinal changes in a multi-modal, multi-scale manner. BNs are probabilistic graphical models, where nodes represent variables and edges represent probabilistic stochastic dependencies between them (12). These stochastic dependencies are characterized by a conditional probability table (CPT) for each variable. These conditional distributions are specified by the network parameters (details in the Supplementary material) (13).

In this work we compiled a BN of 10 risk markers and 10 prodromal markers as well as age. Risk and prodromal markers were selected based on the recent MDS research criteria for prodromal PD, and of which prospective data has been collected in the TREND study. The markers were assigned to different domains including: autonomic dysfunction (constipation, symptomatic orthostatic hypotension, erectile and urinary dysfunction), lifestyle features and related diseases (physical inactivity, non-smoking, diabetes type II), environmental features (occupational pesticide and solvent exposure), neuropsychiatric features (depression, global cognitive deficit), neurological features (incident PD diagnosis (PD conversion based on neurological diagnosis), subthreshold parkinsonism (based on MDS-UPDRS-III), possible REM-sleep behavior disorder (pRBD), hyposmia, substantia nigra (SN) hyperechogenicity), genetic factors (first-degree family history of PD, polygenic risk scores of PD, GBA mutations) and demographic factors (age, sex). Since erectile dysfunction was only assessed in males, this prodromal marker was not included in the final BN to avoid biases to the model. The details of marker assessment methods and definitions are provided in the Supplementary material. This also includes details regarding the handling of missing values.

We employed a BN to learn dependencies between these variables in a data-driven manner as a function of time. BNs result in a quantitative network representing statistical dependencies between variables (12,14). For each variable the probability to take a specific value, dependent on the values of its parents in the network, is inferred from the data. Notably, age (younger or older than 65 years) as well as risk and prodromal markers of PD have been discretized such that all variables indicate the presence or absence (or borderline status) of a marker in an individual TREND participant, as published previously for the TREND cohort (15, 16) and suggested by the MDS research criteria for prodromal PD (3, 6). For each variable a CPT was determined while the overall BN was learned. Conversion to PD was defined as one node in the BN irrespective of the visit at which PD was diagnosed. Further details about the BN learning procedure including the constraints imposed and handling of missing values are reported in the Supplementary material.

We trained a BN based on the data of all 1178 subjects using a non-parametric bootstrap (16) by randomly selecting n = 1178 for 1,000 times, with replacement, and for each of these 1,000 bootstrap samples we learned a complete BN structure. The relative frequency of observing a particular edge (i.e. stochastic dependency) among those 1,000 bootstraps was determined (see BN edges in Figure 1), and served as an indicator of the level of statistical confidence, i.e., a higher value means a stronger support by the data for the existence of the respective connection. A value of 1.0 indicates two specific nodes were interdependent in all the 1,000 learned BNs, a value of 0.5 indicates in 50% of the BNs an interdependency was observed.

**Figure 1:**
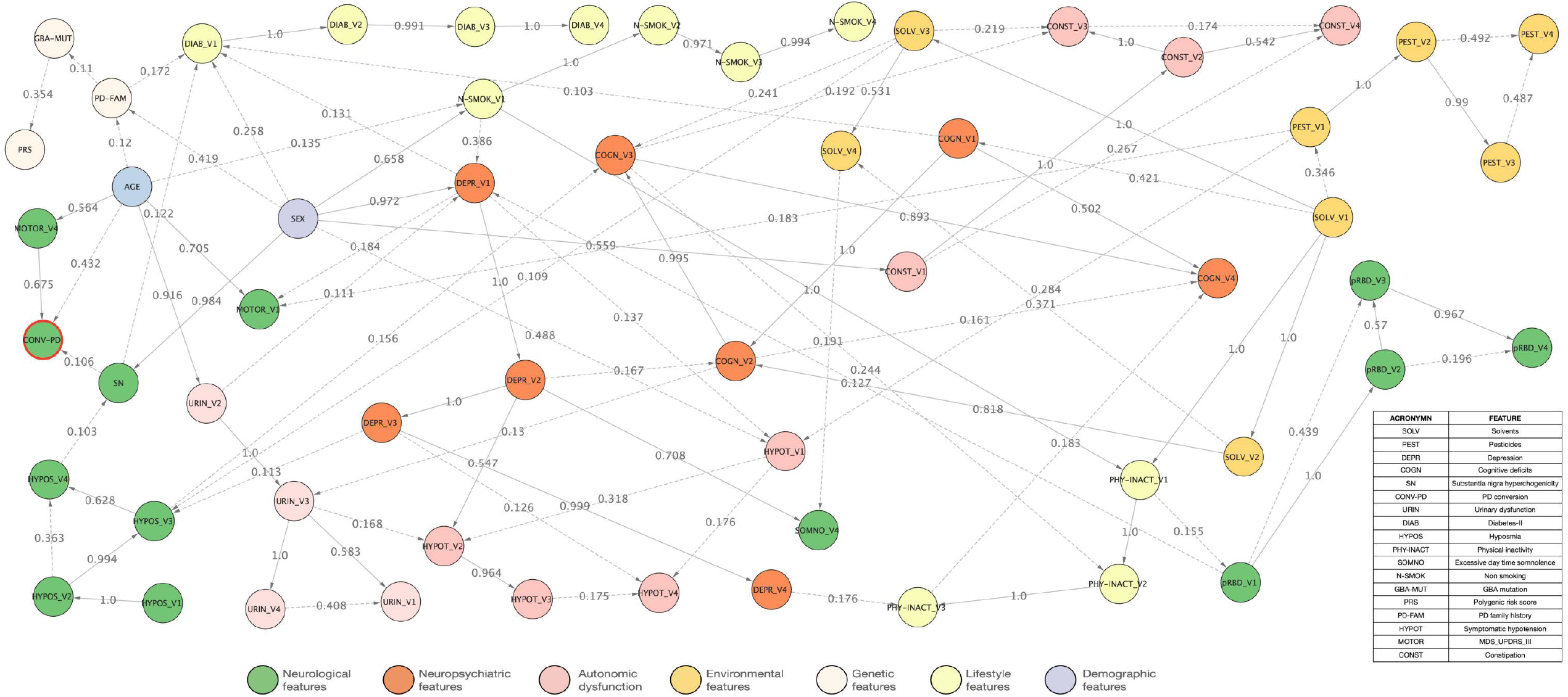
Interdependencies between different risk markers and prodromal markers of Parkinson’s Disease. The depicted Bayesian network represents interdependencies between variables learned from prospective TREND data. Domains of marker nodes are indicated by circle color. The node of the “Conversion to PD” is indicated by a red circle outline. Numbers on edges indicate the level of statistical confidence (bootstrap probability), and dashed edge lines indicate confidences <0.5 while solid lines indicate a confidence ≥0.5. A higher value indicates a higher confidence in the existence of a connection. Nodes isolated from the rest of the network are not shown. V indicates the respective visit number.

### 2.4 Generating synthetic TREND subjects

BNs belong to the class of generative machine learning models. That means they learn the multivariate statistical distribution underlying the observed data. Therefore, random samples drawn from the model correspond to synthetic subjects (see Supplementary material for details). To demonstrate the utility of synthetic subjects generated by the BN model we performed different tests:

1. We generated the same number of synthetic individuals as real individuals for the data and then tested whether a conventional random forest (RF) classifier was able to separate between synthetic and real subjects within 10 times repeated 10-fold cross-validation scheme (5). Therefore, we sequentially left out 1/10 of subjects and trained an RF on the remaining subjects to learn the discrimination between real and synthetic subjects. We used the left-out portion of the data to assess the prediction performance of the RF. We used the partial area under ROC curve (pAUC) at a pre-specified true positive rate of 99% for real subjects as a measure of the prediction performance. The area under the ROC curve at which the detection rate for real subjects was between 99% and 100% served as an indicator of the validity of the synthetic TREND participants. This was done to account for the fact that misclassification of a synthetic TREND participant as real would be far less relevant as the other way around.
2. As a second test we used the synthetic data to learn a BN structure and compared it to the BN learned from real data by counting the fraction of overlapping edges.
3. As a third test, we trained and evaluated the prediction performance of different machine learning models on real as well as synthetic data. More specifically, we here focused on the prodromal markers pRBD, hyposmia and depression. We trained a machine learning model (a random forest classifier) to test the prediction ability of several variables to predict these prodromal markers at multiple visits. Outcomes at a subsequent visit were predicted by training the classifier on variables from the previous visit. For example, to predict the prodromal marker at visit 2, the classifier was trained on all the markers (measured longitudinally in the study) at visit 1. We either trained and tested the classifier on real subjects or trained the classifier on simulated / synthetic subjects generated by the BN and subsequently tested the classifier on real subjects. We evaluated the prediction performance of machine learning models using 10-fold cross validation repeated for 10 times. The overall dataset was randomly split into 10 folds, of which sequentially one of the folds was left out for testing the model, while the rest of the data was used for training. The prediction ability was measured via the area under the receiver operator characteristic curve (AUC) (17).

## 3.0 Results

### 3.1 Descriptive statistics

For each of the four visits, the descriptive statistics of the longitudinal data of risk markers (Table 1) and prodromal markers (Table 2) of PD-free individuals and incident PD cases is shown. Of 1178 subjects, 24 participants were clinically diagnosed with PD over the course of the prospective TREND.

### 3.2 The Bayesian network of risk and prodromal markers of PD in the TREND study

Figure 1 depicts the overall network structure of all connections learned from the TREND data. A wide range in the level of confidence regarding the interconnectedness, i.e., statistical interdependence, was observed between several nodes and domain clusters of nodes. High probabilistic confidence (>0.5) of edges between different markers in the BN was found for edges between age to subthreshold parkinsonism (MDS-UPDRS-III) and urinary dysfunction, sex to SN hyperechogenicity, depression, non-smoking and to constipation; depression to symptomatic hypotension and excessive daytime somnolence; solvent exposure to cognitive deficits and to physical inactivity; and non-smoking to physical inactivity. Pairwise co-occurrences of different markers showing edges with probabilistic certainties of >0.2 in the BN were shown and statistically tested for significance in Table 3. All of these edges also showed statistically significant co-occurrences between markers, except for sex and PD family history, sex and diabetes type-II (visit 1), occupational solvent exposure (visit 3) and constipation (visit 3), as well as GBA mutation carriers and PRS. These associations were no longer significant after accounting for multiple testing.

**Table 3:** Statistical testing of the co-occurrence of risk and prodromal marker pairs (A & B) in the TREND data (including imputed data) as suggested by edges in the TREND BN of real data. P-values have been calculated based on a *χ*^2^-test and corrected for multiple testing using Holm’s method. Significant findings (after Holm-Bonferroni correction for multiple testing) are indicated by an asterisk. Findings remain significant in logistic regressions additionally accounting for age and sex. V, visit.

| Risk/prodromal marker A | Risk/prodromal marker B | Risk/prodromal marker B<br>Participants (n) |  |  | p-value |
| --- | --- | --- | --- | --- | --- |
|  |  | No | Borderline | Yes |  |
| Male sex | Depression (V1) | 471 |  | 128 | <0.0001* |
| Female sex |  | 359 |  | 220 |  |
| Male sex | Non-smoker (V1) | 228 | 320 | 51 | <0.0001* |
| Female sex |  | 308 | 213 | 58 |  |
| Male sex | SN hyperechogenicity | 453 |  | 146 | <0.0001* |
| Female sex |  | 515 |  | 64 |  |
| Male sex | Constipation (V1) | 549 | 45 | 5 | <0.0001* |
| Female sex |  | 472 | 96 | 11 |  |
| Male sex | PD family history | 529 |  | 70 | 0.013 |
| Female sex |  | 481 |  | 98 |  |
| Male sex | Symptomatic hypotension (V1) | 508 | 83 | 8 | <0.0001* |
| Female sex |  | 412 | 147 | 20 |  |
| Male sex | Diabetes type II (V1) | 567 |  | 32 | 0.024 |
| Female sex |  | 564 |  | 15 |  |
| Exposure to solvents (V1) | Cognitive deficits (V1) | 302 |  | 86 | <0.0001* |
| No exposure to solvents (V1) |  | 678 |  | 112 |  |
| Exposure to solvents (V2) | Cognitive deficits (V2) | 246 |  | 176 | <0.0001* |
| No exposure to solvents (V2) |  | 682 |  | 74 |  |
| Exposure to solvents (V3) | Cognitive deficits (V3) | 201 |  | 237 | <0.0001* |
| No exposure to solvents (V3) |  | 668 |  | 72 |  |
| Exposure to solvents (V3) | Constipation (V3) | 394 | 32 | 12 | 0.029 |
| No exposure to solvents (V3) |  | 626 | 88 | 26 |  |
| Exposure to pesticides (V1) | Symptomatic hypotension (V1) | 13 | 20 | 1 | <0.0001* |
| No exposure to pesticides (V1) |  | 907 | 210 | 27 |  |
| Presence of depression (V2) | Day time somnolence (V4) | 414 |  | 26 | <0.0001* |
| Absence of depression (V2) |  | 725 |  | 13 |  |
| Presence of depression (V2) | Symptomatic hypotension (V2) | 213 | 200 | 27 | <0.0001* |
| Absence of depression (V2) |  | 588 | 122 | 28 |  |
| Non-smoker at visit (V1) | Physically active (V1) | 107 |  | 429 | <0.0001* |
| Borderline smoker (V1) |  | 82 |  | 451 |  |
| Smoker (V1) |  | 63 |  | 46 |  |
| Non-smoker (V1) | Depression (V1) | 389 |  | 147 | <0.0001* |
| Borderline smoker (V1) |  | 382 |  | 151 |  |
| Smoker (V1) |  | 59 |  | 50 |  |
| Presence of Global cognitive deficits (V2) | Physically active (V3) | 224 |  | 85 | <0.0001* |
| Absence of Global cognitive deficits (V2) |  | 193 |  | 676 |  |
| GBA mutation carriers | Polygenic risk score | 21 | 24 | 7 | 0.002 |
| GBA mutation non-carriers |  | 226 | 655 | 245 |  |
| Age (> 65 years) | Conversion to PD | 500 |  | 23 | <0.0001* |
| Age (≤ 65 years) |  | 654 |  | 1 |  |
| Age (> 65 years) | Subthreshold parkinsonism (V4) | 462 | 31 | 30 | <0.0001* |
| Age (≤ 65 years) |  | 616 | 32 | 7 |  |
| Age (> 65 years) | Subthreshold parkinsonism (V1) | 418 | 71 | 34 | <0.0001* |
| Age (≤ 65 years) |  | 588 | 49 | 18 |  |
| Age (> 65 years) | Urinary Dysfunction (V1) | 269 | 220 | 34 | <0.0001* |
| Age (≤ 65 years) |  | 465 | 173 | 17 |  |
| Age (> 65 years) | Non-smoking (V1) | 260 | 236 | 27 | <0.0001* |
| Age (≤ 65 years) |  | 276 | 297 | 82 |  |

The BN revealed both expected as well as novel connections between risk and prodromal markers and the phenoconversion to PD. Plausibly, the nodes with edges directed to the conversion to PD comprised (prior) subthreshold parkinsonism indicated by MDS-UPDRS-III scores, age, and (with lower statistical confidence), SN hyperechogenicity. Further expected marker interdependencies were observed for edges pointing from depression and solvent exposure to global cognitive deficits, which itself was linked to physical inactivity while non-smoking was linked to physical inactivity. Edges pointing from depression to excessive daytime somnolence, pointing from solvent exposure and depression to hyposmia, or pointing from hyposmia to global cognitive deficits and to SN hyperechogenicity demonstrated further expected interdependencies. Unexpected interdependencies were observed from depression to non-smoking; pesticide exposure to symptomatic hypotension; physical inactivity to urinary dysfunction; and edges with directionality from SN hyperechogenicity, global cognitive deficits, sex and PD family history to diabetes. Interestingly, constipation was dependent on sex, global cognitive and occupational solvent exposure. Surprisingly, little interdependencies were observed for pRBD, which was only linked to depression and received an edge from physical inactivity. Nodes with genetic features were not dependent on other markers except for sex being linked to PD family history, which itself was linked to diabetes.

Nodes of the same marker assessed at different timepoints were largely highly interdependent, except for subthreshold parkinsonism (MDS-UPDRS-III) for which visit 2 and visit 3, which were not linked to other nodes of the BN. MDS-UPDRS-III at visit 1 showed no edge with the corresponding nodes of other visits, but instead only received edges from depression and pesticide exposure at visit 1. An interactive Cytoscape network file of the BN is given in the Supplementary material.

### 3.3. Simulation of a synthetic TREND study cohort

The generative property of the BN allowed the simulation of synthetic versions of the prospective data of the TREND study and to extract individual synthetic participant profiles including age and the risk and prodromal markers of PD. Table 4 shows five arbitrary examples of synthetic subjects (from the synthetic cohort with the same sample size) and three real subjects together with their individual data (at visit 4) on age, sex, MDS-UPDRS-III, pRBD, depression, global cognitive deficits and PD conversion status. The Multiple Correspondence Analysis (MCA) (18) plot shown in Figure 2 indicates the similarity of synthetic subjects in relation to real ones. Further systematic comparisons of the distribution of individual variables and their correlation structure are presented in the Supplementary material (Figures S2-S6). An RF classifier trained to discriminate between real and synthetic subjects only performed slightly better than chance level (pAUC 52%), indicating that both real and synthetic subjects cannot be reliably discriminated (Figure S7).

**Table 4.**
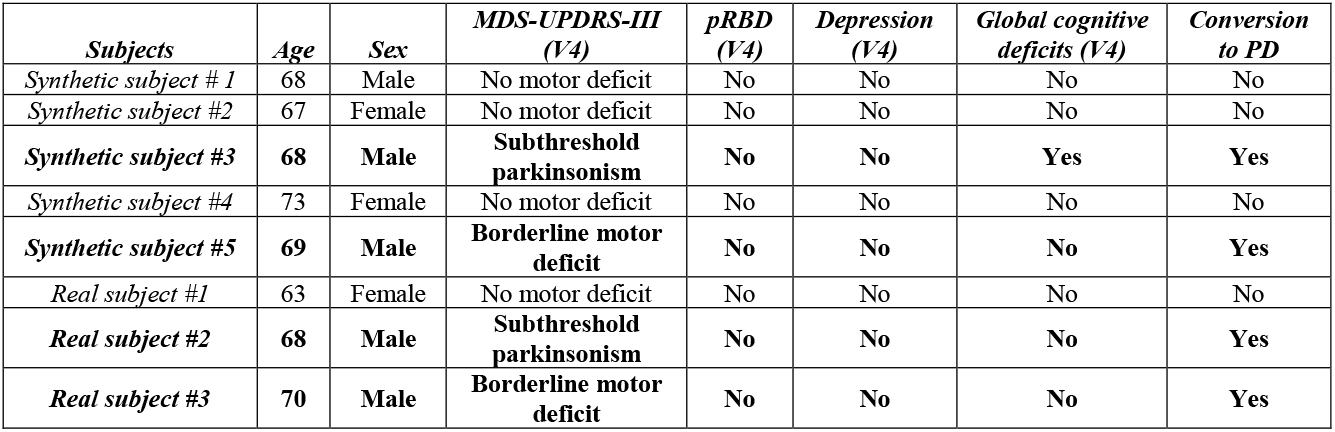
Examples of synthetic and real subjects and their demographics, selected prodromal markers, subthreshold parkinsonism (MDS-UPDRS-III) and PD conversion status at visit 4. The rows in bold represent the similarity between the real and synthetic subjects’ data for incident PD cases. MDS-UPDRS-III, subthreshold parkinsonism indicated by the MDS-sponsored Unified Parkinson’s Disease Rating Scale; PD, Parkinson’s Disease, pRBD, possible REM-sleep behavior disorder. V, visit.

| *Subjects* | *Age* | *Sex* | *MDS-UPDRS-III (V4)* | *pRBD (V4)* | *Depression (V4)* | *Global cognitive deficits (V4)* | *Conversion to PD* |
| --- | --- | --- | --- | --- | --- | --- | --- |
| *Synthetic subject #1* | 68 | Male | No motor deficit | No | No | No | No |
| *Synthetic subject #2* | 67 | Female | No motor deficit | No | No | No | No |
| ***Synthetic subject #3*** | **68** | **Male** | **Subthreshold parkinsonism** | **No** | **No** | **Yes** | **Yes** |
| *Synthetic subject #4* | 73 | Female | No motor deficit | No | No | No | No |
| ***Synthetic subject #5*** | **69** | **Male** | **Borderline motor deficit** | **No** | **No** | **No** | **Yes** |
| *Real subject #1* | 63 | Female | No motor deficit | No | No | No | No |
| ***Real subject #2*** | **68** | **Male** | **Subthreshold parkinsonism** | **No** | **No** | **No** | **Yes** |
| ***Real subject #3*** | **70** | **Male** | **Borderline motor deficit** | **No** | **No** | **No** | **Yes** |

**Figure 2.**
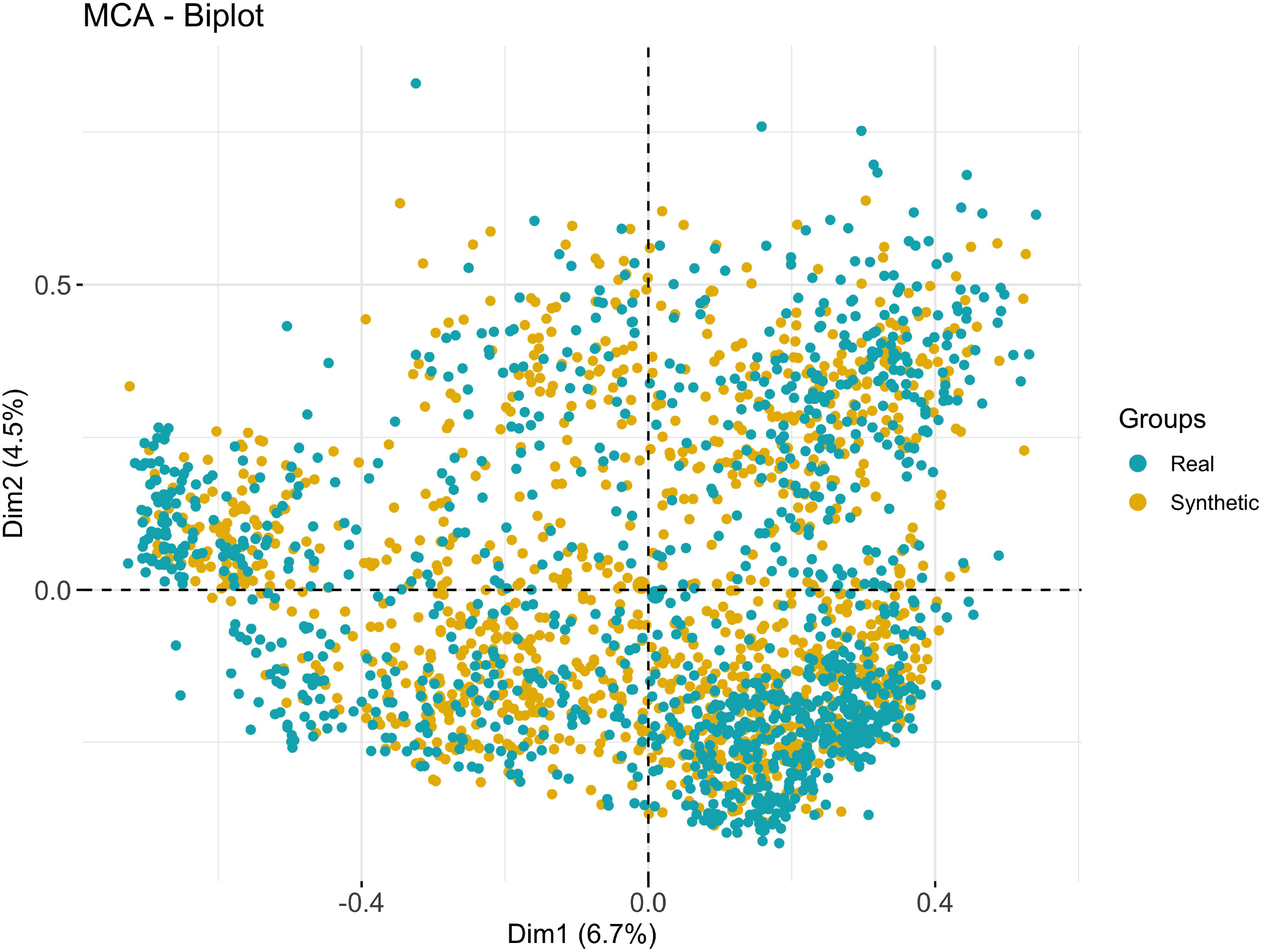
Multiple correspondence (MCA) analysis plot of prospective data of real (in blue) and simulated (in yellow) TREND participants.

### 3.4 Evaluating the utility of synthetic TREND subjects

We repeated the training of the BN based on the synthetically generated data. Subsequently, we counted, how many robust marker edges (bootstrap probability >50%) were common between the original BN trained on real and the BN trained on synthetic data. Accordingly, we could establish a high degree of similarity of 84.12% (143 edges) between the BNs learned from synthetic and real data. To further evaluate the utility of synthetically generated TREND subjects we developed RF classifiers to predict for the individual participant, whether a participant would develop pRBD, hyposmia and/or depression at subsequent visits of the study. As outlined in the Methods part of this paper, corresponding classifiers were trained within a 10-times repeated 10-fold cross-validation, once on real subjects and once on synthetically generated subjects. To account for the possible variability due to the random sampling of synthetic subjects from the BN model, the process was repeated 10 times. Models were always tested on real patients.

Despite synthetic data generally showing a high similarity to real data, our results indicate a loss of ∼10% AUC when training on synthetic compared to training on real subjects (Figure S9). This could be due to slight differences between real and synthetic data regarding the distribution of individual variables (e.g. hyposmia, physical inactivity, see Figure S4) as well as correlation structure (Figure S6). Notably, RFs are a comparably complex machine learning method, which allows for modeling highly nonlinear structures.

Altogether these results highlight that synthetic data share many patterns of real patient data, but they are not identical and hence do not necessarily allow for coming to identical statistical conclusions.

## 4.0 Discussion

The present study shows the feasibility of generating a Bayesian network based on prospective data of established risk and prodromal markers of PD in the TREND cohort of elderly PD-free individuals and incident PD cases: (1) The BN model showed several expected as well as unexpected interdependencies of these markers, which may be explained by biological and clinical reasons for the co-occurrences of markers and/or by confounding due to practical and other methodological aspects of marker assessment. (2) The BN allowed to create a synthetic representation of the TREND cohort regarding marker interdependencies and to derive realistic marker profiles of individual synthetic participants. The multitude of marker interdependencies as revealed through the artificial intelligence-based BN modelling approach could have important methodological implications for evidence-based PD prediction approaches as well as for the understanding of the interplay of different markers in the prodromal phase of PD.

The current methodological approach of the MDS research criteria for prodromal PD (20,21) uses a naïve Bayesian classifier for the prediction of PD (or diagnosis of prodromal PD), which assumes that predictive values of risk and prodromal markers are independent. Based on our findings from a BN model and pair-wise testing of co-occurrences of established PD markers in the prospective TREND cohort, we could show that for many of these predictive markers the assumption of statistical independence is most likely not met. Hence, concerns about the validity of the naïve Bayes classifier approach for PD prediction are raised.

While the number of incident PD cases was relatively low in the present study, robust and plausible interdependency was observed between prior motor deficits indicating subthreshold parkinsonism and the phenoconversion to PD. Also, age and SN hyperechogenicity were linked to the incidence of PD, which was expected as the prevalence of PD markedly increases with advancing age (20,21) and SN hyperechogenicity is observed in 83% of PD patients (22). However, for SN hyperechogenicity a substantially lower certainty was present in the bootstrapping of the BN models as may be partly explained both by low number of incident PD cases and by potential prodromal differences between distinct subtypes of the disease, e.g., the hypothesized brain-first vs. body-first prodromal PD subtypes (23–25). Among risk and prodromal markers of PD, which have been shown to also play a role in other neurodegenerative and neuropsychiatric conditions, several interdependencies were observed in the TREND BN model. Depression is a risk factor of cognitive decline but may also affect motivational aspects reducing cognitive performance in challenging neuropsychological test situations, and both may underlie the interdependency between depression and global cognitive deficits (26,27). Occupational solvent exposure has been associated with an increased risk of global cognitive impairment (28), which is consistent with their observed interdependency in the BN. Global cognitive deficits were linked to physical inactivity, which is an established risk factor for cognitive decline and dementia (29). As expected, current smokers were less physically active than former smokers and non-smokers explaining the edge between non-smoking and physical inactivity. Similarly, non-smoking was linked to depression and smokers were more frequently depressed than non-smokers. Given the known protective effects of smoking for PD (30) and increased PD risk due to physical inactivity and depression (31,32), these often co-occurring factors may have opposing effects for individual PD risk estimates. Excessive daytime somnolence has been shown to be both a risk factor for depression as well as a frequent comorbid factor in depressed individuals, supporting their interdependency observed in the BN (33). Depression has been previously linked to lower olfactory performance and, conversely, patients with olfactory dysfunction have been reported to show symptoms of depression that worsened with the severity of smell loss (34). Hyposmia has been shown to be a risk factor for cognitive impairment and neurodegeneration (35). However, odor discrimination tests might be more cognitively challenging in those with cognitive deficits (36), which might in part also explain the interdependency between hyposmia and global cognitive deficits. Olfactory loss is observed in the vast majority of PD patients (37), and as SN hyperechogenicity is also highly prevalent in PD patients these markers may plausibly frequently co-occur in clinical as well as prodromal PD. However, only a low level of confidence of this edge was observed in the BN, and the interdependency of hyposmia and SN hyperechogenicity may be more robust in specifically selected individuals in the prodrome of PD, possibly in particular in brain-first prodromal PD.

Occupational pesticide exposure was interdependent with symptomatic hypotension, however both markers have been assessed without highly specific methods. The degree and specific exposure to pesticides might vary widely among the TREND participants, and symptomatic hypotension has been assessed via self-report questionnaire only, and thus may not constitute neurogenic orthostatic hypotension (20). The statistical confidence was not high for the interdependency between these markers and thus remains to be further investigated. Sedentary lifestyle has been shown to increase the risk of urinary incontinence (38,39), which might underlie the edge between physical inactivity to urinary dysfunction. Moreover, age showed a robust edge to urinary dysfunction, which is plausible as this prodromal marker becomes more prevalent with aging. Diabetes received interesting edges from several nodes including SN hyperechogenicity, global cognitive deficits, sex and PD family history, and while their confidences were low, this finding might provide new hypotheses regarding biological prodromal mechanisms to be tested in future studies.

Several node interdependencies were unexpected and should be further investigated in independent cohorts. Possible RBD was only assessed using a self-report questionnaire, and while we applied the most specific criteria to determine the presence and absence of (possible) RBD (40), polysomnography would likely reveal a high false-positive rate among pRBD as a prevalence of polysomnography-proven RBD is less than 2% in the general, elderly population (41). Low specificity of the assessment methods might have contributed to the lack of interdependencies between possible RBD and many other risk and prodromal markers of PD, including markers of autonomous dysfunction, which, together with RBD, may often co-occur in a body-first prodromal PD subtype (23). pRBD was linked to depression and physical inactivity, which, however, might in part correspond to unspecific sleep disturbances in depression (42) and (obese) physically inactive individuals (e.g., with obstructive sleep apnea) (43). Interestingly, constipation (a hypothesized body-first prodromal PD marker) showed interdependency with global cognitive deficits and occupational solvent exposure (both hypothesized brain-first markers), but not with other autonomous dysfunctions or pRBD. Possibly, as constipation is frequent in old age, its specificity for (incident) PD and thus interdependence with other risk/prodromal markers of (body-first) PD might be low. However, the relationship between constipation and other risk/prodromal markers of PD and their collective predictive values for PD (subtypes), and, e.g., PD risk conveyed through the gut microbiome (44), should be further investigated. Genetic risk markers of PD were, except for sex and diabetes, not interdependent with other risk and prodromal markers of PD, and while the number of GBA mutation carriers was low, a positive PD family history and a high polygenic risk score may increase the PD risk in a highly complex and multifaceted manner, which may partly explain the lack of their direct interdependency with other risk and prodromal markers. While we expected age to be interdependent with several other markers frequent in old age (e.g., constipation, SN hyperechogenicity, hyposmia, global cognitive deficits), yet such edges were not observed in the BN and accounting for age did largely not alter effects of pair-wise co-occurrences in our logistic regression analysis. As expected, nodes of the same marker assessed at different visits were largely highly interdependent. Subthreshold parkinsonism based on MDS-UPDRS-III at visit 1 however showed no edge with the corresponding nodes of other visits, and the data of visits 2 and 3 were interdependent with one another, yet not connected to the BN. Possibly, motor deficits were either not (yet) apparent in some participants or motor deficits may have been confounded with non-PD related arthritic, tendon, bone or muscle complications at the first visit.

BN-based sharing of cohort information in prodromal PD research might be a means to provide researchers with a “preview” on patient-level data without having to go through a typically time-consuming legal process, which is necessary to get access to real data. While our analysis has shown that synthetic data shares many characteristics of real data, it is not identical and therefore statistical analysis results could differ between real and synthetic data. The intent behind the generation of synthetic data is thus not to replace the analysis of real data, but to facilitate the understanding of a data source by data analysts in a simple manner. Notably, the Dutch Netherlands Cancer Institute (NKI) has, based on similar considerations, recently launched a synthetic cancer cohort on their webpage: https://iknl.nl/en/ncr/synthetic-dataset. A similar strategy might be useful for the prodromal PD research community to facilitate data sharing and to overcome the still existing data silos. In this context, it should be emphasized that one of the key bottlenecks in modern medicine is the fact that clinical studies are scattered across organizations. Each clinical study is unavoidably biased by inclusion and exclusion criteria, therefore raising concerns about the representativeness of a particular study for the overall (prodromal) disease population. It is thus necessary that results obtained with data from one study are cross-checked with those from another one to generate sufficient evidence, and this is in particular true for machine learning models. Comprehensive cohort data of population-based studies often cannot be shared due to legal restrictions. Therefore, techniques are required, which lower the hurdle for a legally compliant sharing of clinical and population-based cohort data. While federated data analysis techniques are now gaining a lot of interest, setting up an according network of organizations is highly challenging from a technical as well as management point of view. Synthetic data could fill a gap, because organizations are potentially willing to share such data more easily. Still, appropriate protection measures for synthetic data should be taken to avoid the theoretically still existing possibility of adversarial attacks (19).

The present study has several limitations that need to be discussed. 1. Despite an excellent retention rate in the TREND study participant attrition as well as missing data for single visits were observed in the longitudinal data. While we comprehensively imputed missing data while accounting for data missing not at-random, it might have introduced slight biases into the data and marker interdependencies. 2. Inter-visit dependency of markers, such as ratings of motor deficits, might in part be lowered due to changes of investigators between different waves of TREND data collection and assessment. 3. While a directionality of edges between markers is proposed by our BN approach, alternative directions may be observed in different models. However, indeed directions not predefined by constraints were largely expected. 4. BN structure and parameter learning requires sufficiently large datasets that are representative for the disease population. The BN model thus renders the re-identification of real patients from the training data relatively unlikely. However, in its current implementation our approach does not provide strict theoretical guarantees for this situation. But we like to point out that privacy preserving training of neural network models is possible and has been tested by us and others in the past (45).

### 4.1 Conclusion

In conclusion, the present study used a BN to disentangle the relationships of various established risk and prodromal markers in a large prospective cohort and showed that many of these markers are interdependent. Interdependencies of these predictive markers have not been accounted for in current PD prediction approaches, such as the MDS research criteria for prodromal PD (20,21). Moreover, the independent predictive value of individual markers has not been determined or incorporated into PD prediction models as comprehensive marker data of prospective cohorts is hardly published or accessible. While the BN of the TREND cohort contained data of a large sample of PD-free individuals, yet only a small sample of incident PD cases were available. Hence, an accurate PD prediction accounting for the interdependencies in marker profiles could not be derived from the given data.

We demonstrated that the BN model of TREND cohort data could be used to generate a sufficiently realistic cohort of synthetic TREND subjects, which would allow researchers to obtain a useful “preview” on TREND data. This has to be seen in the context of the fact that PD prediction accounting for the heterogeneity, complexity and temporal dynamics of (prodromal) PD (subtypes) will require further collaboration and data sharing in the future, e.g., by jointly investigating and integrating data (or models of these data) across multiple observational cohorts with a large cumulative number of incident PD cases. In addition, the current trend regarding the analysis of large real-world data sources (e.g., electronic health records) should not be ignored.

Overall, this work demonstrates the potential of modern AI approaches to advance our understanding of prodromal PD and to facilitate data sharing.

## Data Availability

All data produced in the present study are available upon reasonable request to the authors

## Acknowledgments

We thank the participants for their continued participation in the TREND study and for providing their biosamples; the numerous (doctoral) students and study nurses, who actively contributed to study organization, and data collection, entry, and monitoring.

## Author Contributions

Contributed to the conception and design of the study: HF, SH; Contributed to implementation of methods, data analysis and running the experiments: MS; Contributed to drafting the text, preparing the figures: MS, HUZ, SH, HF; Contributed to the conception of the TREND cohort study, TREND supervision and data acquisition: US, A-KT, KB, GWE, WM, DB; All authors have read and approved the manuscript in its current form.

## Data Availability Statement

The TREND cohort data can be shared upon request.

