## Supplementary Material for "Bayesian network modeling of risk and prodromal markers of Parkinson’s disease"

The ten risk markers and ten prodromal markers of Parkinson's disease (PD) as assessed in the TREND study were defined as reported previously (1) and selected according to the recent International Parkinson and Movement Disorder Society (MDS) Research Criteria for Prodromal PD (2). The assessment methods of the risk and prodromal markers of PD and the coding of the presence, absence and borderline status, respectively, as considered in the Bayesian Network modelling are described below.

**Risk markers of PD**

- **Sex:** male=0, female=1
- **Smoking status:** self-report questionnaire; categorized into 0 = never smoker; 1 = former smoker; 2 = current smoker
- **Family history of PD:** self-report questionnaire; any first degree relative with PD; no=0 yes=1.
- **Polygenic PD risk:** determined based polygenic risk scores calculated using data of 90 single nucleotide polymorphisms according to Nalls et al. (3); lower quartile of the TREND PRS distribution = 0; upper quartile = 1 (higher polygenic risk); borderline = 2 (quartile 2 and 3))
- **Glucocerebrosidase (GBA) gene mutations:** Presence of GBA het T369M (p.T408M), GBA het T297S (p.T336S), GBA het N370S (p.N409S), GBA het E326K (p.E365K); no=0 yes=1.
- **Hyperechogenicity of the substantia nigra (SN+, SN- status):** assessed using transcranial B-mode sonography (TCS) following the standard imaging protocols. In case the bone window did not allow the measurement of the SN the data was indicated as missing. One SN of at least one hemispheric side larger than the designated cut-off value of 0.23 $cm^2$ was considered abnormal (4). No (SN-) = 0, Yes (SN+) = 1.
- **Occupational pesticide exposure:** self-report questionnaire on occupational pesticide exposure; no=0 yes=1.
- **Occupational solvent exposure:** self-report questionnaire on occupational solvent exposure; no=0 yes=1.
- **Diabetes type II:** defined as being treated with antidiabetic medication (insulin or oral antidiabetics); no=0 yes=1.
- **Physical inactivity:** standardized questionnaire (5); less than one hour of exercise per week or no exercise in the last three months was considered physical inactivity; no=0 yes=1.

**Prodromal markers of PD:**

- **Possible rapid eye movement sleep behavior disorder (pRBD)**: pRBD has not been assessed using polysomnography (hence possible RBD) but based on the RBD screening questionnaire (RBDSQ), and the criteria suggested by Marelli et al. (6) were considered for the definition of possible RBD (cut-off=8 without item number 10). Participants who scored at least 8 in the questionnaire were considered to have an pRBD; no=0 yes=1.
- **Subthreshold parkinsonism:** assessed using the Movement Disorder Society-unified Parkinson's disease rating scale, motor part III, (MDS-UPDRS-III), and defined as MDS-UPDRS-III total scores > 6 excluding postural and action tremor items (7). Individuals with MDS-UPDRS-III < 3 were considered negative. Individuals between 3-6 in the MDS-UPDRS-III were considered borderline; no=0, borderline=1, yes=2.
- **Hyposmia:** assessed using the Sniffin' Sticks odor identification test (SS-16, Burghart Medizintechnik, Germany). Normative criteria for age and sex were applied according to

Hummel et al.(8). Individuals with abnormal olfaction were given LR+=6.4, whereas normosmia had LR-=0.40. In case of severe cold, allergies, or any other medical reason for abnormal olfaction, LR=1 was considered. assessed by the Sniffin' Sticks odor identification (SS-16 version; Burghart Medizintechnik, Wedel, Germany).

- **Depression:** assessed using a self-reported medical diagnosis of depression. Additionally, the presence of an episode of major depression according to ICD-10 criteria was assessed by the Major Depression Inventory (MDI) (9). Individuals were considered for scoring positive for depression if they had a lifetime history of depression or were currently suffering from acute depression (0= no depression; 1=acute/life-time depression.
- **Autonomic dysfunctions** were assessed by the UMSARS questionnaire (Unified Multiple System Atrophy Rating Scale, Part I7) (10).
- **Constipation:** Individuals with a score of at least 2 (Frequent constipation requiring the use of laxatives) were considered having constipation. Individuals with a score of 1 (Occasional constipation but no medication needed) were considered borderline. Participants with scored 0 (No change in bowel function pattern from the previous pattern) were considered to have no constipation; no=0, borderline=1, yes=2.
- **Symptomatic hypotension:** A score of at least 2 (Orthostatic symptoms developing at least once a week) were considered abnormal. A score of 1 "Orthostatic symptoms are infrequent and do not restrict activities of daily living) was considered borderline, and a score of 0 (No orthostatic symptoms) indicated the absence of the marker; no=0, borderline=1, yes=2.
- **Urinary dysfunction:** A score of at least 2 (Urgency and/or frequency, drug treatment required) defined the presence of an abnormality. A score of 1 (Urgency and/or frequency, no drug treatment required) indicated a borderline case, and the score of 0 was considered normal; no=0, borderline=1, yes=2.
- **Erectile dysfunction:** all assessed using conservative marker definitions applying the Unified Multiple System Atrophy Rating Scalewith a cut-off of 2 for constipation, symptomatic hypotension (orthostasis). and urinary dysfunction, and of 3 for erectile dysfunction (Scores of 1 or 2 (Minor or Moderate impairment compared to healthy days) were considered borderline); no=0, borderline=1, yes=2.
- **Global cognitive deficit:** assessed using the battery of "Consortium to Establish a Registry for Alzheimer's Disease" neuropsychological battery (CERAD) (11). Individuals who scored worse than -1.0 standard deviation in at least two out of four domains were considered positive for the global cognitive deficit; no=0 yes=1.

### Bayesian Networks

#### General

Bayesian Networks (BN) are probabilistic graphical models, where nodes represent variables and edges represent probabilistic stochastic dependencies between nodes. A BN for a set of variables X = $X_1$,....$X_n$ is a network structure S that encodes a joint statistical distribution of X by factorizing it according to a given directed acyclic graph (DAG) G = V,E into local conditional distributions. Here nodes correspond V to the variables in X and arcs E represent direct dependencies between variables. The variables in the form of nodes are represented by $X_i$ and $Pa_i$ denotes the parents of node $X_i$ in S and the variables corresponding to those parents. According to Markov chain rule; where every random variable $X_i$ is directly dependent on its parents $Pa(x_i)$ (12) the joint probability distribution for all the variables represented by a BN can be decomposed into a product of conditional probabilities using the graphical structure and the chain rule of probability calculus:

$$p(x|\theta) = \prod_{i=1}^{n} p(x_i|pa(x_i), \theta_i)$$

where $x_i = x_1, x_n$ are the variables (nodes in the BN) and $\theta = \theta_1,\ldots,\theta_n$ are the BN's parameters, where each $\theta_i$ defines the set of parameters that are important to define the distribution of the variables given its parents $pa(x_i)$. The terms in the product correspond to the local probability distributions P and joint distribution p(x) is represented by the pair (S, P).

In data with longitudinal measurements, such as in the TREND study, there exists a subset $X' \subset X$ in which measurements are time dependent, i.e. x' = (x'(1))…..x'(T)) with T being the number of visits. Dynamic Bayesian Networks (13) usually account for the time-dependency of measurements by implicitly unfolding the BN structure over time, i.e. introducing for each visit t, a separate copy X'(t) of X' while requiring that edges always point from time slice t to time slice t + 1 (corresponding to a first order Markov process). This implicit unfolding assumes a stationary Markov process, i.e. parameters θ do not change with time. In our setting this assumption is most likely wrong, because individuals change in their marker profile and disease outcome (incident PD) during the course of a study, i.e. $p(X'(t)|X'(t-1)) \neq (X'(t+1)|X'(t))$. Hence, we here use an unfolding strategy, in which we explicitly use different copies X'(t) for each time point.

**Handling missing values**

One of the key challenges with longitudinal data and especially the data that is missing not at random (MNAR) (14). Missingness both due to study drop-out (until visit 4) as well as due to missingness of marker assessment at single visits was considered for the imputation of data. We mitigated this challenge by defining auxiliary variables for each feature that has been measured longitudinally, as described in our earlier publication (15). The auxiliary variables were defined for each variable at each visit. Thereafter we used the missForest (16) method to impute the missing values. For the features not having longitudinal measurements, we imputed the missing values using the most frequent label for that feature as all of them were factor variables. Later, for the BN we also created a common missing feature at every visit to account for higher order of missingness.

**Structure and parameter learning of the BN, including non-parametric bootstrap**

Structure learning consists of finding the optimal directed acyclic graph and parameter learning involves describing the conditional probability distributions (17). The problem of learning the structure of BN from a complete dataset D that comprises d data points, $D = D_1, D_2, \ldots D_d$ corresponds to determine the set of directed arcs E for DAG G = (X,E) using a criterion that specifies the fit of the structure to the observed data, in our case the Bayesian Information Criterion (BIC). The process of structure learning is NP hard (18) and a substantial amount of work in the research community has been dedicated to identify heuristic search techniques to identify good models. As the number of potential graphs increases exponentially with the number of variables (19), identification of the true data generating graph structure is computationally challenging. In this context it is important to mention that - under the assumption of no unknown statistical confounders – the true causal graph structure is known to be a part of a class of equivalent graph structures, called class partially directed acyclic graph (CPDAG) (20). Considering this assumption, the CPDAG has the same structure as the true causal graph but might also contain undirected edges. Therefore, it is important to restrict the CPDAG equivalence class as much as possible by prior knowledge to allow correct directionality of as many edges as possible. For this purpose, we imposed the following constraints for the BN structure:

- No node can affect environmental factors except the factors itself
- Genetic factors like GBA mutation and Polygenic risk score (PRS) cannot be dependent on any other feature except themselves, PD family history and sex
- PD family history cannot be dependent on any other features except genetic factor, age and sex
- No other feature can be affected by conversion feature (Conversion node is the node that represents if the subject has converted to PD or not)
- Age and gender cannot be dependent on any other node
- Auxiliary variable that was created based on missingness of a certain feature can only have an influence on their corresponding feature and the auxiliary variable for the same feature at the next time point

We learned the BN using four different algorithms (hill climbing, tabu search, max-min hill climbing, 2-phase Restricted Maximization) from R-package bnlearn (21). K-fold cross-validation was used to assess the generalization ability of a BN model and compare different structure learning algorithms. That means the overall data was randomly split into k (k = 10) folds, and the BN structure together with its parameters was learned using k-1 folds. If the overall population is correctly modeled by the fitted BN (and not just the training data), the data in the left-out fold with high probability should fall into the same statistical distribution that is described by the BN. In practice the negative expected log-likelihood of the test data was used to quantify the validation.

Accordingly, we selected a hill climbing (hc) search for subsequently learning a final BN from the data (Figure S1) (22). At this point we employed a non-parametric bootstrap to quantify the confidence in the existence of each edge. That means we re-sampled for 1000 times d data points with replacement and based on each bootstrap sample a BN structure was learned using hc search.

After identification of a BN structure, parameters (i.e. conditional probability tables) were inferred using a Dirichlet prior to account for variable configurations in parents of a specific node that could not be observed in the data.

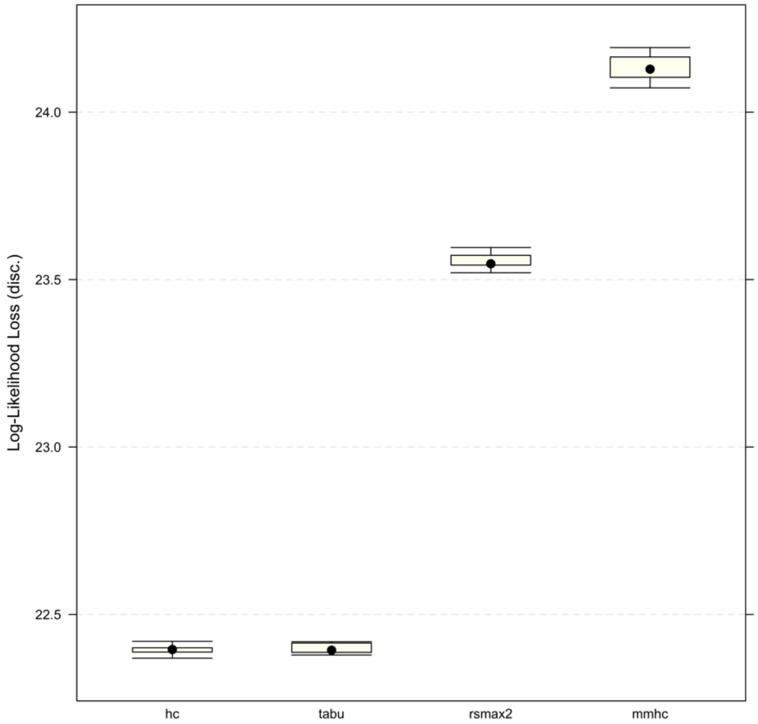


Figure S1. Comparison of different BN structure learning algorithms via 10-fold cross-validation for the data. The y-axis depicts the negative log-likelihood of the test data.

### Simulation of synthetic subjects

We exploited the fact that BNs belong to the class of generative models. That means they describe a multivariate distribution, from which it is possible to sample synthetic subjects. This was done by first drawing random values from a node's distributions and subsequently from the distributions of the children of that node while conditioning on the values of the parents. We sampled synthetic subjects as real participants and then compared the distributions of each variable visually at each visit (Figure S2, S3, S4, S5) as well as by calculating the Kullback-Leibler divergence (KLD) (23). KLD is used to quantify the distributions of a random variable and measure the similarity between the two probability distributions of the same variable. Hence, it measures the divergence of one distribution from another. If the distributions are an exact match, the KL divergence is 0, otherwise it can lie between 0 and ∞.

As seen in the underlying figures, the KLD value is close to 0 that means we have a good match between the distribution of real and synthetic data, the lower the value the better is the match.

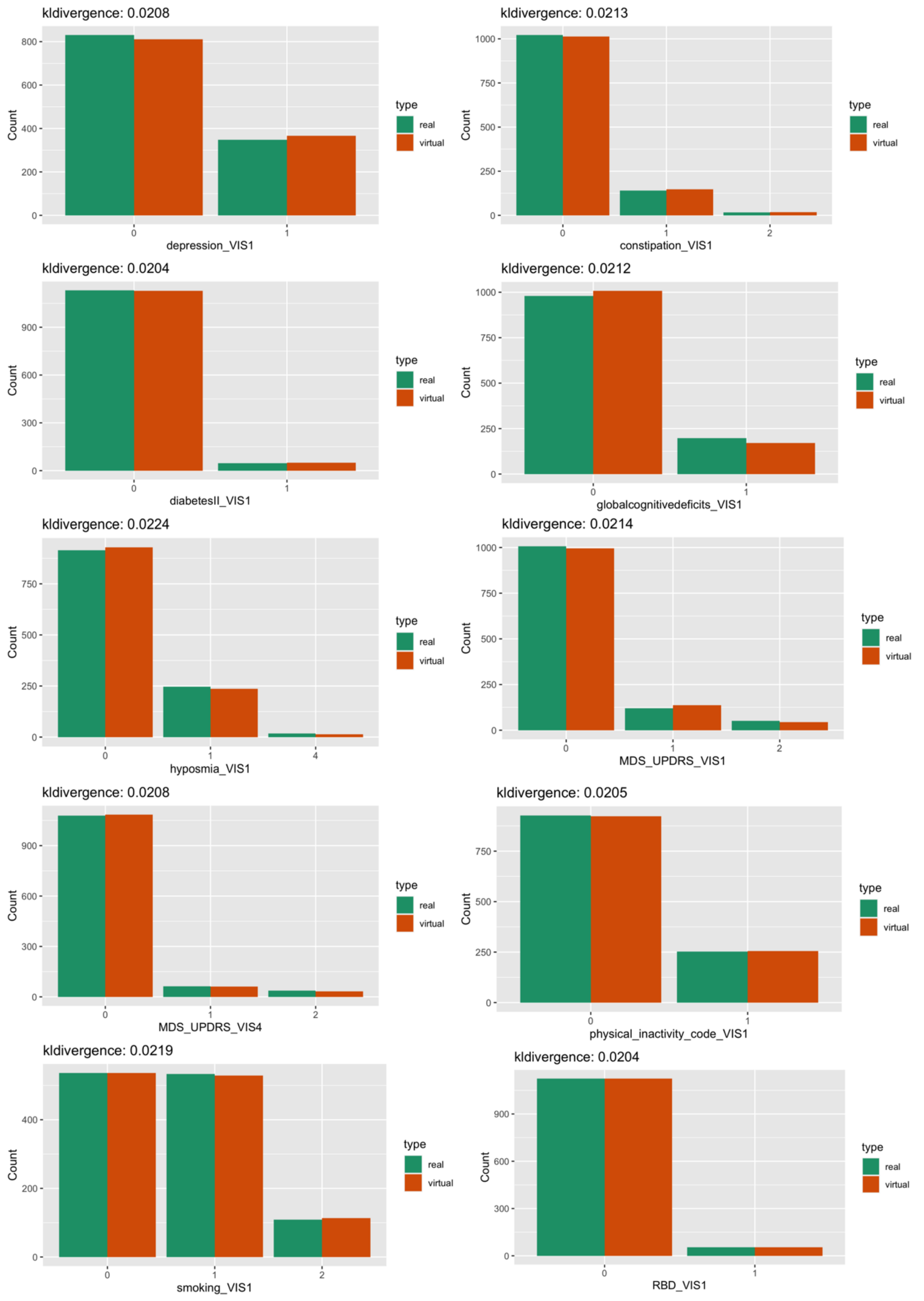


Figure S2. Examples of real and simulated subjects at visit 1 generated via the BN model trained on TREND study data. The Figure compares the distributions of features at visit 1 for real subjects (green) and synthetic / simulated subjects (red). KL-divergence between the real and synthetic subjects is mentioned on the top of each plot.

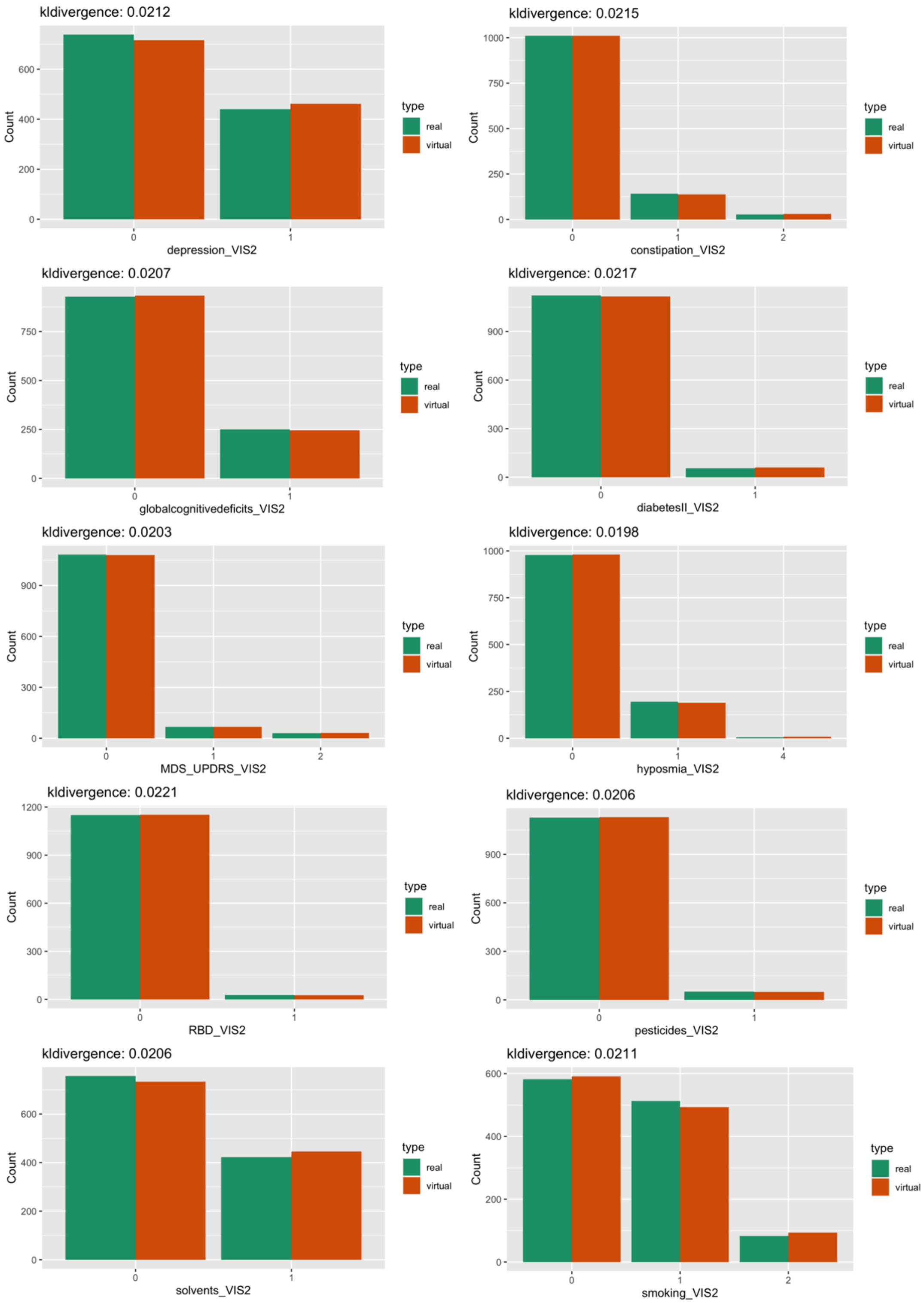


Figure S3. Examples of real and simulated subjects at visit 2 generated via the BN model trained on TREND data. The Figure compares the distributions of features at visit 2 for real subjects (green) and synthetic / simulated subjects (red). KL-divergence between the real and synthetic subjects is mentioned on the top of each plot.

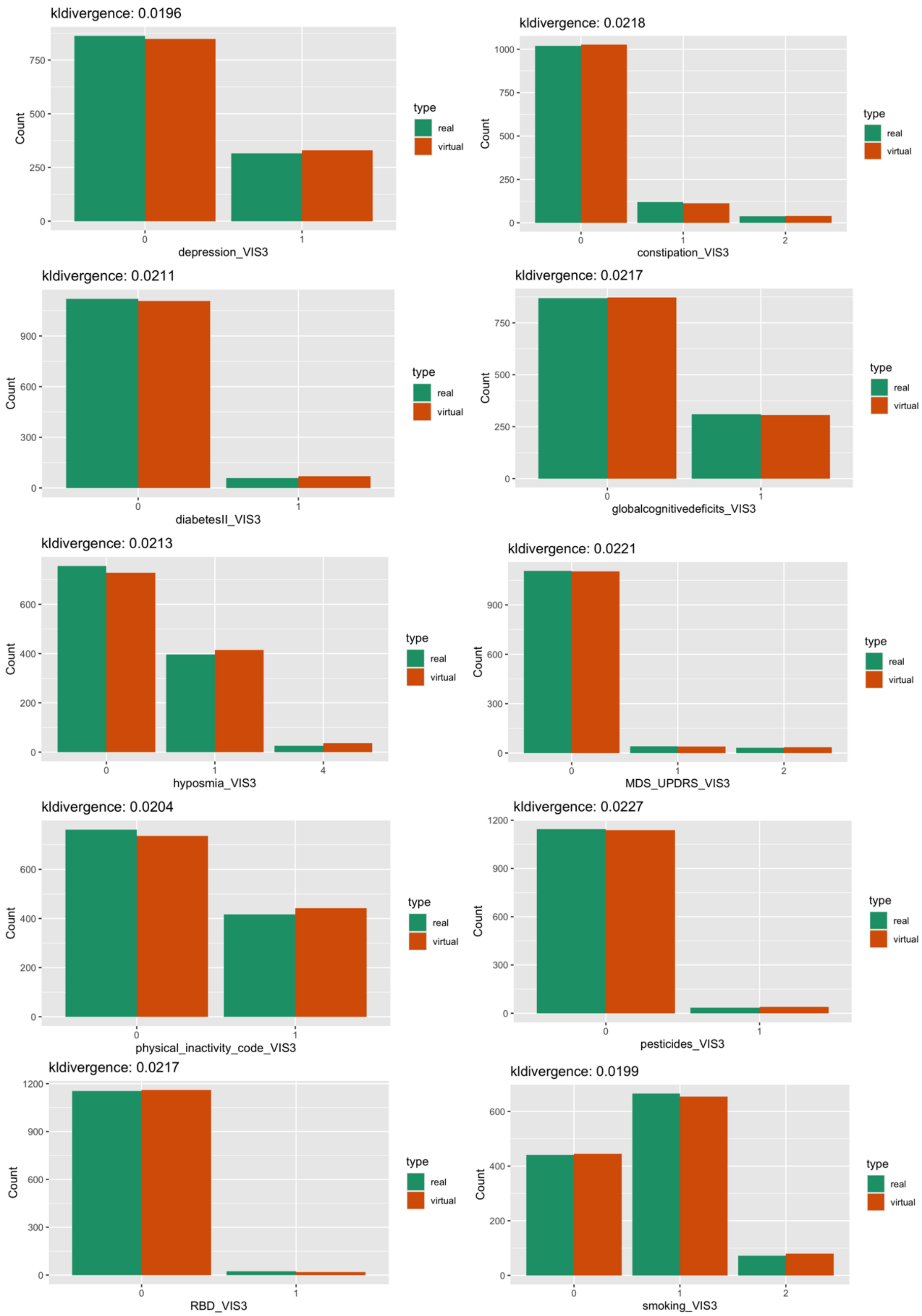


Figure S4. Examples of real and simulated subjects at visit 3 generated via the BN model trained on TREND data. The Figure compares the distributions of features at visit 3 for real subjects (green) and synthetic / simulated subjects (red). KL-divergence between the real and synthetic subjects is mentioned on the top of each plot.

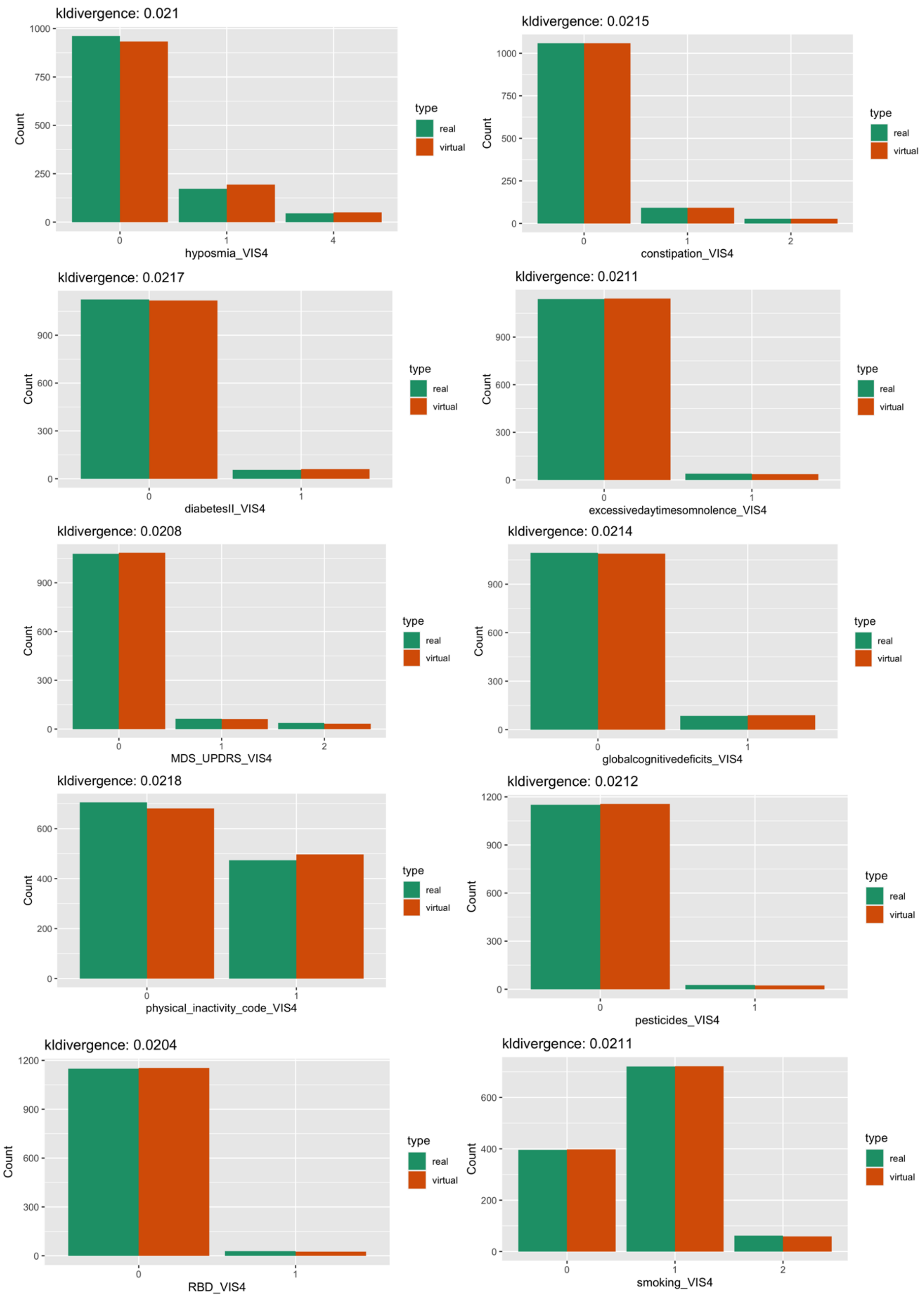


Figure S5. Examples of real and simulated subjects at visit 4 generated via the BN model trained on TREND data. The Figure compares the distributions of features at visit 4 for real subjects (green) and synthetic / simulated subjects (red). KL-divergence between the real and synthetic subjects is mentioned on the top of each plot.

We also measured spearman rank correlation for real and synthetic data, and we observe that the correlation between the variables remain preserved. The plot is illustrated in Figure S6.

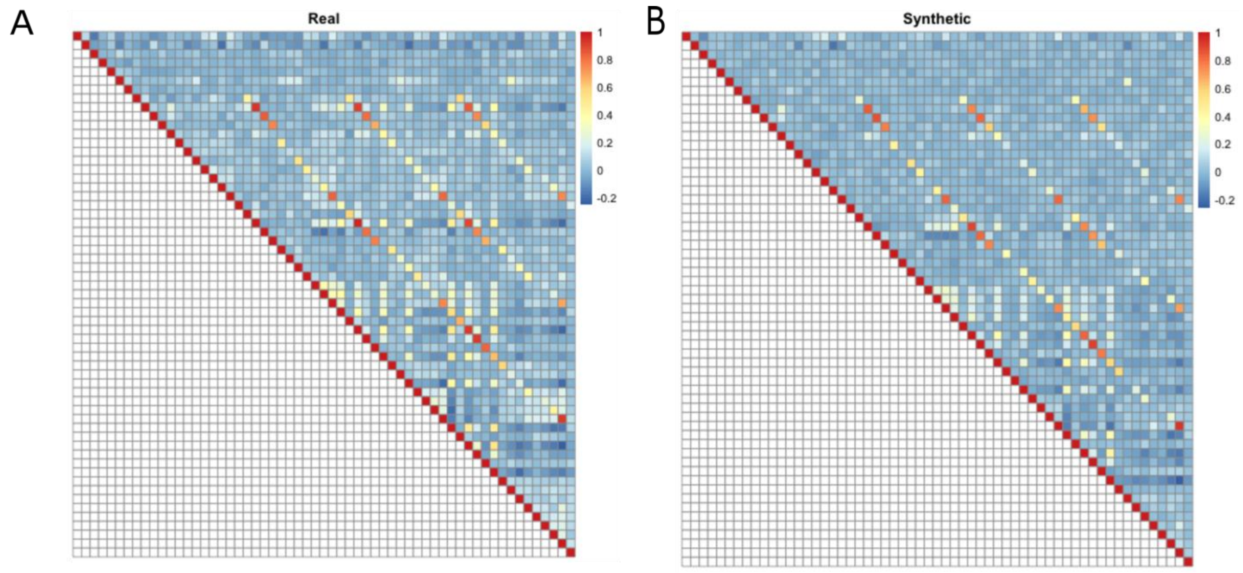


Figure S6. Spearman rank correlation for (A) real and (B) synthetic subjects.Legend indicates the strength of correlation, the darker the color (red or blue) the higher is the correlation (positive (red) or negative(blue))

### Testing the validity of our synthetic TREND participants

In order to validate our synthetic TREND participants, we generated the same number of synthetic individuals as real individuals for the data and then tested whether a conventional RF classifier was able to separate between synthetic and real subjects within 10 times repeated 10-fold cross-validation scheme (5). That means we sequentially left out 1/10 of subjects and trained a RF on the remaining subjects to learn the discrimination between real and synthetic subjects. We used the left-out portion of the data to assess the prediction performance of the RF. We used the partial area under ROC curve (pAUC) at a pre-specified true positive rate of 99% for real subjects as a measure of the prediction performance. The area under the ROC curve at which the detection rate for real subjects was between 99% and 100% served as an indicator of the validity of the synthetic TREND participants. This was done to account for the fact that misclassification of a synthetic TREND participant as real would be far less relevant as the other way around. In our case a pAUC only slightly above chance level was achieved (Figure S7, indicating that synthetic subjects cannot reliably be discriminated from real ones by machine learning.

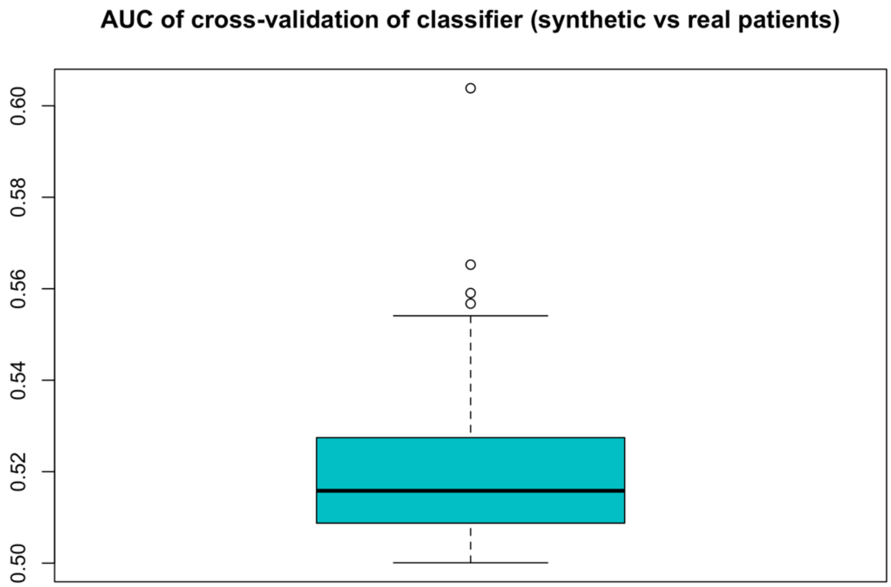


Figure S7. Performance of a random forest classifier to correctly identify a given number of real TREND participants among synthetic subjects. The performance was measured via the partial area under ROC curve (pAUC) at a pre-specified detection rate of ≥ 99% for real participants. The pAUC was assessed on test sets within 10 repeats of a 10-fold cross-validation procedure. Accordingly, boxplots show the distribution of the tenfold cross-validated pAUC that was obtained from 10 repeats of the cross-validation procedure.

The RF method assigns to each synthetic subject a confidence score, namely the probability to belong to the class of real subjects. Figure S8 visualizes these confidence scores with a color code in a multiple correspondence analysis plot (a technique similar to PCA that is devoted to discrete data (24)).

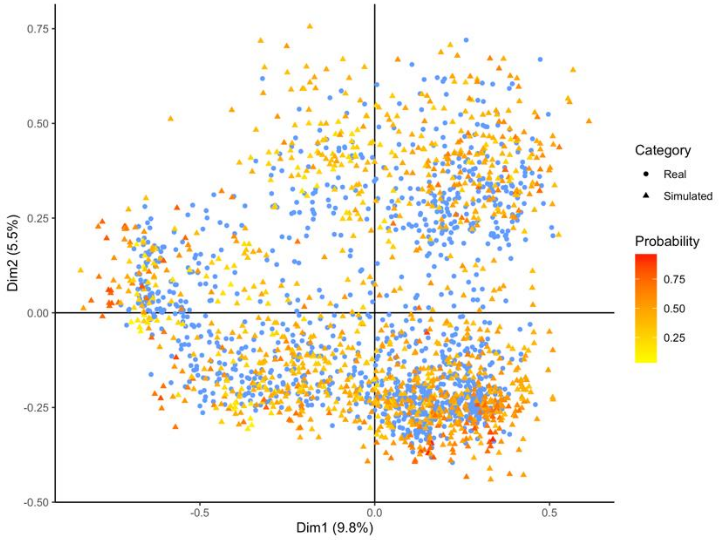


Figure S8. Multiple correspondence analysis plot of real ADNI and synthetic subjects. The probability of a synthetic participant to belong to the real data is indicated by color.

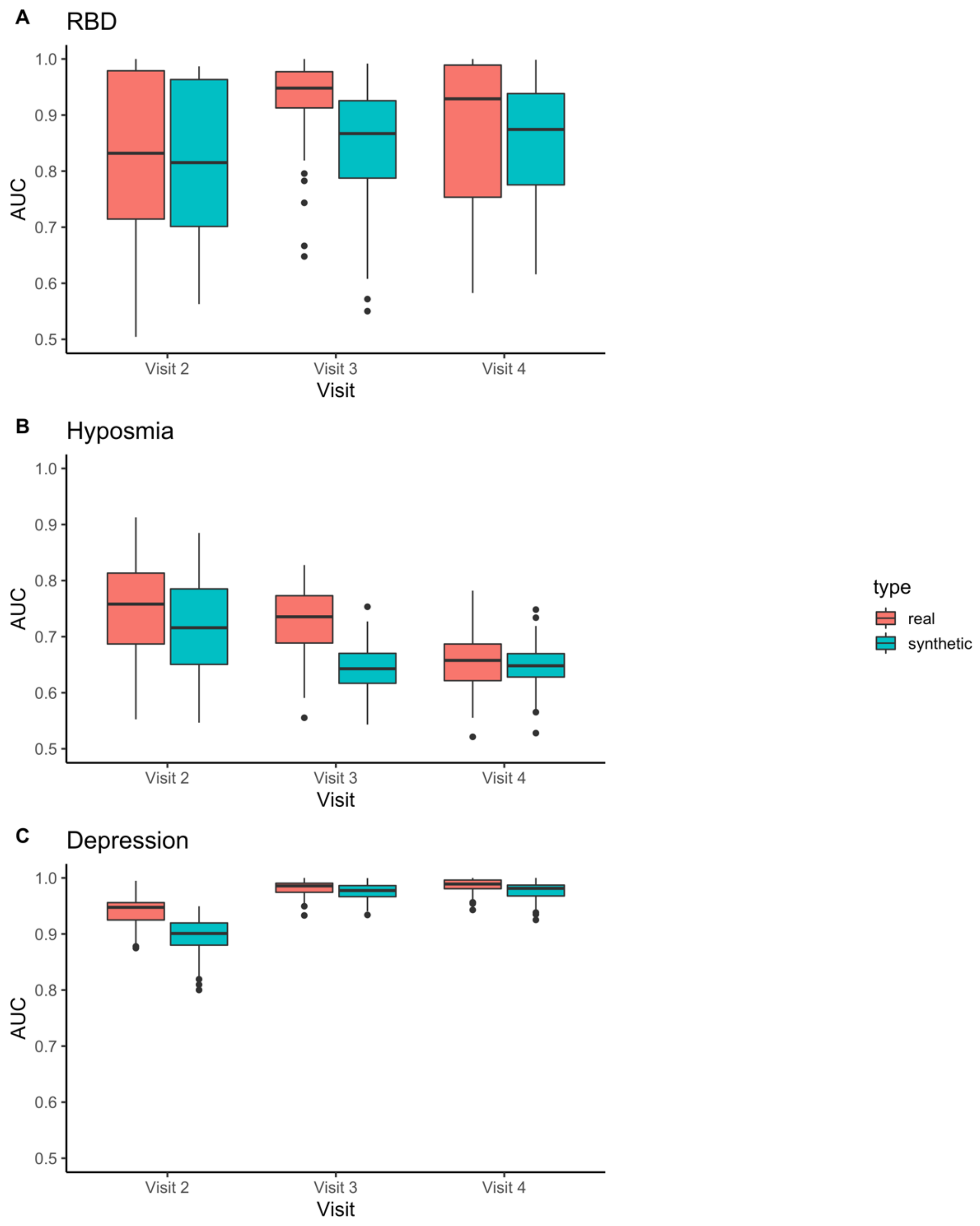


Figure S9. Performance of a random forest classifier with subjects trained on real and tested on real data (red), trained on synthetic and tested on real data (green), and trained on synthetic and tested on synthetic data (blue) for **(A)** pRBD **(B)** hyposmia and **(C)** depression as clinical end points of the prospective data. The classification accuracy is indicated as area under the curve (AUC) in Receiver-Operating-Characteristic (ROC) curves. The boxplots show the distribution of the AUC derived from the real/synthetic training data within 10 times repeated 10-fold cross-validation. In case of training on synthetic data, results have been averaged over 50 repeated samplings of the same number of synthetic as real subjects.
